# Buying time: an ecological survival analysis of COVID-19 spread based on the gravity model

**DOI:** 10.1101/2020.05.01.20087569

**Authors:** Alon Vigdorovits

## Abstract

COVID-19 has spread in a matter of months to most countries in the world. Various social and economic factors determine the time in which a pandemic reaches a country. This time is essential, because it allows countries to prepare their response. This study considered a gravity model that expressed time to first case as a function of multiple socio-economic factors. First, Kaplan-Meier analysis was performed for each variable in the model by dividing countries into two groups according to the median of the respective variable. In order to measure the effect of these variables, parameters of the gravity model were estimated using accelerated failure time (AFT) survival analysis. In the Kaplan-Meier analysis the differences between high and low value groups were significant for every variable except population. The AFT analysis determined that increased personal freedom had the largest effect on lowering the survival time, controlling for detection capacity. Higher GDP per capita and a larger population also reduced survival time, while a greater distance from the outbreak source increased it. Understanding the influence of factors affecting time to index case can help us understand disease spread in the early stages of a pandemic.

## 1. Introduction

COVID-19 is the disease caused by the novel SARS-CoV-2. It has rapidly spread across the world and developed into a pandemic. By the 23^rd^ of April, there have been over 2.5 million cases since the disease was first reported in Wuhan, China, on the 31^st^ of December (ECDC, 2020b; Rothan & Byrareddy, 2020).

The transmission of an infectious agent depends on various factors. Freedom of movement shapes the course of an outbreak, and governments can influence freedom directly by enacting a state of emergency (Nay, 2020). Urbanization creates clusters of high population density where respiratory pathogens can spread easily (Eisenberg et al., 2007). The increased accessibility of air travel has also accelerated disease transmission (Mangili et al., 2015). Increased economic activity facilitates human contact, leading to increased transmission (Tatem et al., 2006). Analysis of COVID-19 spread from one country to another would benefit from accounting for these factors.

The gravity model is a framework borrowed from transportation theory. It can also be used to model the spread of infectious diseases (Kraemer et al., 2019). In 2011, the gravity model was validated on data from the 2009 A (H1N1) pandemic (Li et al., 2011).

In this paper, I propose a heuristic form of the gravity model which considers a number of variables potentially associated with country to country spread of COVID-19. The objective of the study is to describe the influence of these variables on the amount of time a country has to prepare for the arrival of a pandemic, once the pandemic has already started. This effect on time to first case is quantified using survival analysis.

## 2. Methods

### 2.1 Gravity model

A gravity model for the intensity of spread can be written as follows (Viboud et al., 2006):

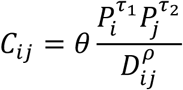

where *C_ij_* is the intensity of spread between communities *i* and *j* of populations *P_i_* and *P_j_*, and *D_ij_* is the distance between communities. The parameters *θ*, *τ*_1_, *τ*_2_ and *ρ* are to be estimated. Greater intensity of spread leads to a faster propagation of the disease to neighboring communities (Li et al., 2011). This means shorter periods of time until the neighboring communities experience their first case. In addition to population size and distance, economic and political factors can also influence spread. Thus, I propose the following model:

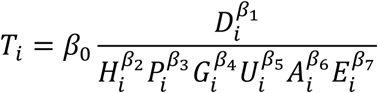

Where *T_i_* is the number of days until the index case in country *i*, *D_i_* is the distance from country *i* to the country where the disease originated (China), *H_i_* is a measure of human and economic freedom in country *i*, *P_i_* is the population, *G_i_* is the GDP per capita, *U_i_* is the degree of urbanization, *A_i_* is the volume of air travel, and *E_i_* is the epidemiological detection capacity. *β*_0_, *β*_1_, *β*_2_, *β*_3_*, β*_4_, *β*_5_, *β*_6_, *β*_7_ are the model parameters. The model can be rewritten by taking the logarithm of both sides:

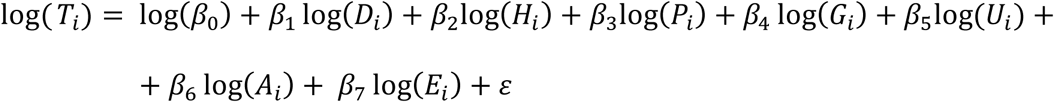

If we let log(*β*_0_) = *µ*; log(*D_i_*) = *X*_1_; *log*(*H_i_*) = *X_2_* and so on for all variables, we obtain:

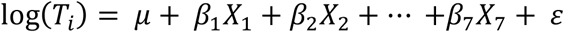

This equation is the log-linear representation of the accelerated failure time (AFT) model, which is a parametric survival model (Wei, 1992).

### 2.2 Data sources

Time from the beginning of the outbreak to the first case in each country was gathered from ECDC public data on COVID-19 on the 11^th^ of April (ECDC, 2020a). The 2019 Human Freedom Index (HFI) was used as a measure for *H_i_* (Cato Institute, 2019). GDP per capita, urbanization percentage, and air transportation (total passengers carried) data were obtained from the latest available World Bank datasets (World Bank, n.d.). Measures for detection and reporting capacity were extracted from the 2019 Global Health Security Index (Johns Hopkins Center for Health Security, 2019). Distances between centroids of countries and China were calculated using the R package geosphere (Robert J. Hijmans, 2019).

### 2.3. Statistical analysis

A total of 156 countries were considered for the analysis, all of them experiencing their COVID-19 case by the 11^th^ of April 2020. Five countries without air travel data were treated as missing at random and dropped from the analysis. China, as the starting point, was not included. The starting date of the analysis was considered the 30^th^ of December 2019. Median, minimum and maximum survival times (times to first case) were determined. Each independent variable in the model was divided by its median into two groups. The survival probability of each group was then assessed using Kaplan-Meier estimates. The survival probabilities of the “low” (below median) and “high” (above median) groups were compared using the log-rank test. P-values were considered significant below 0.05. The variables (continuous, in log form) were included in the AFT model, to evaluate their individual effect on survival time. The best fitting distribution for the AFT model was chosen using Akaike’s Information Criterion (AIC) (Bozdogan, 1987). P-values of model coefficients were considered significant below 0.05. A second model was designed, with the HFI separated into its constituent parts, personal freedom and economic freedom. Cox proportional hazards regression was also performed, and the proportional hazards assumption was tested using Schoenfeld’s residuals (Grambsch & Therneau, 1994). Statistical analysis was performed in R.

## 3. Results

The first countries affected had either geographical proximity to China or very high economic development. Developing countries reported cases later, particularly those at a considerable distance from China. Figure 1 depicts survival times across the globe.

**Figure 1:**
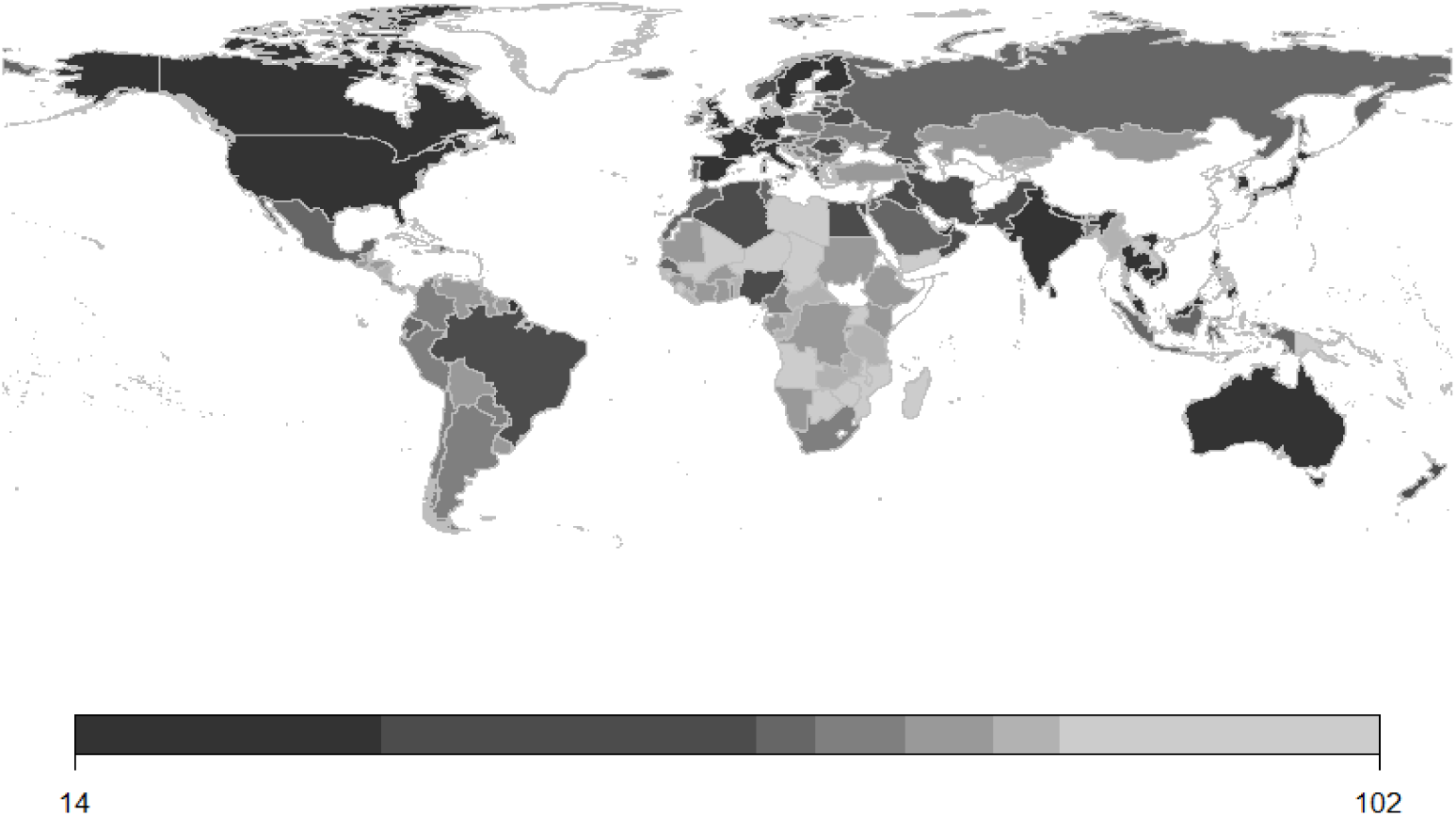
Number of days to first case

Median time to first case was 68 days. The first affected country (after China) was Thailand, at 14 days. The last affected was Yemen at 102 days. As Figure 2 shows, the distribution of survival Median time to first case was 68 days. The first affected country (after China) was Thailand, at 14 days. The last affected was Yemen at 102 days. The distribution of survival times is bimodal, with a group of neighboring and developed countries reporting their first cases in the first wave. The rest of the world was affected in a larger, second wave. Between 35 and 52 days only two countries reported their index case.

**Figure 2:**
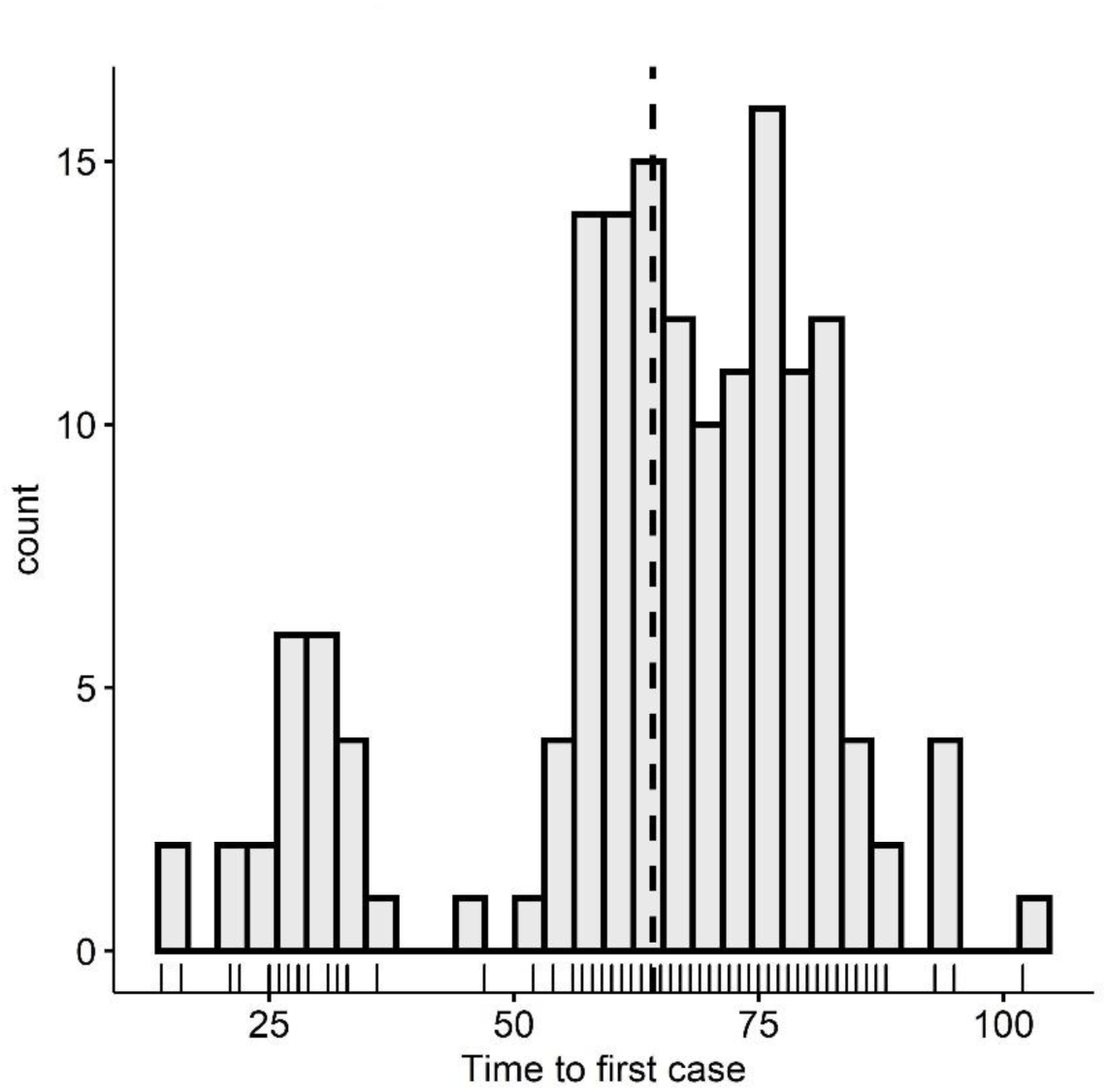
Distribution of survival times (days)

The survival curve for all countries is shown on the top left of Figure 3. The period in which few countries reported index cases is the flat portion of the survival curve. The independent variables of the model, each divided into two groups by their median, are depicted in Figure 3. The log-rank test showed that the observed difference in survival between “low” and “high” groups is statistically significant (p < 0.0001) for all variables except population (p = 0.49). Longer distance from China was associated with longer time to event. For the other variables, lower values were associated with longer time to event.

**Figure 3:**
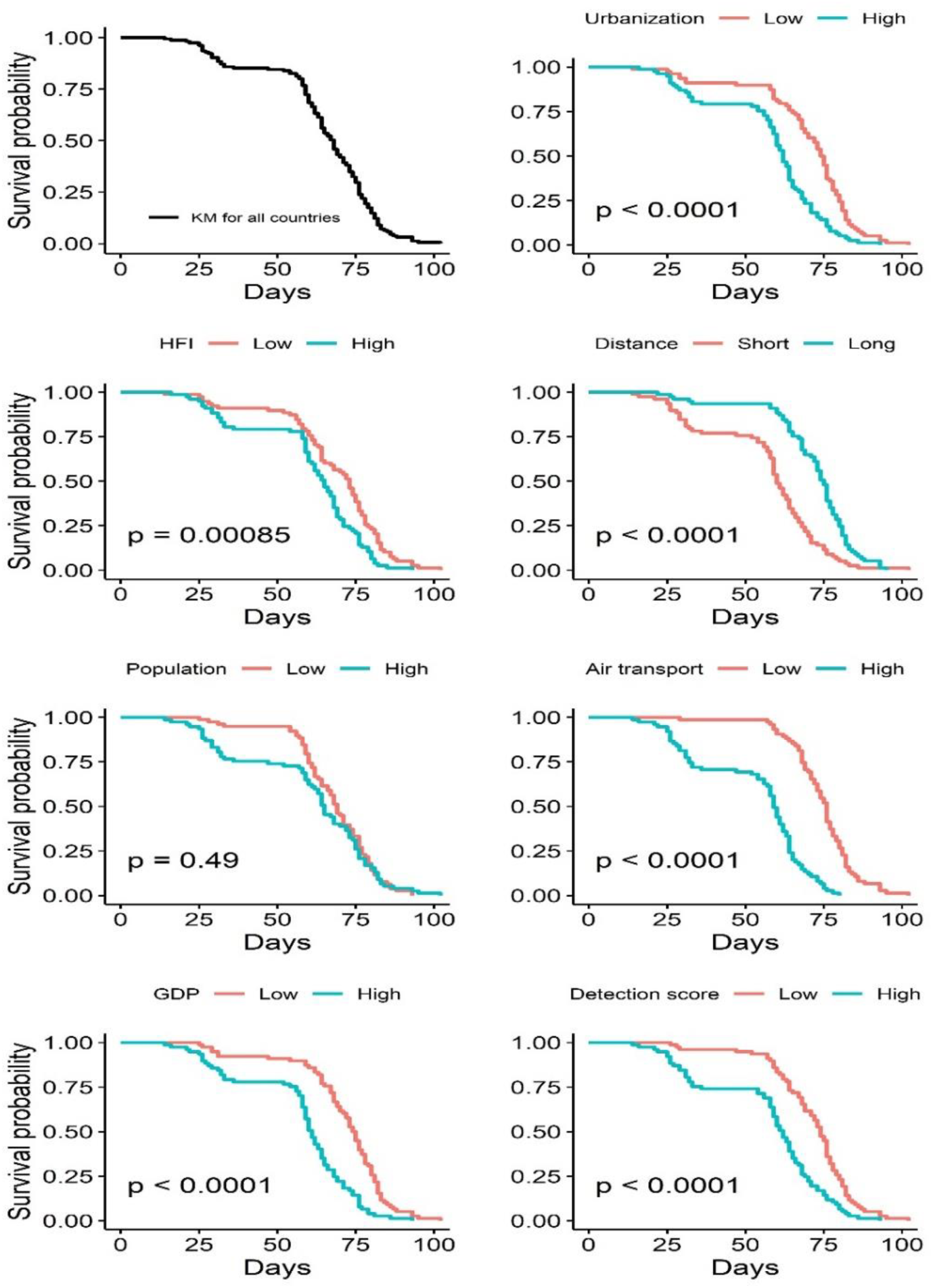
Overall Kaplan-Meier curve (top-left) and Kaplan-Meier curves comparing upper and lower value groups for each variable in the model.

Variables were then included in the AFT model. Based on the AIC, the best distribution to fit the data was the Gompertz distribution. As shown in Table 1, HFI, population, and GDP per capita were significantly associated with survival time (p < 0.05). The model is in log-log form, which means that a variable coefficient is interpreted as the percentage change in survival time given a 1% change in the variable, or the elasticity of the survival time with respect to the variable. Coefficients were positive for all variables except distance. HFI had the largest effect, with a coefficient of 2.46. The weakest effect is that of air transportation volume, with a coefficient of 0.058.

**Table 1 -.** AFT model with Human Freedom Index.

| **Variables** | **Coefficient <sup>†</sup>** | **95 % C.I.** | **P-value** |
| --- | --- | --- | --- |
| Human Freedom Index | -2.46 | (-4.01, -0.91) | 0.002 |
| Population | -0.348 | (-0.51, -0.19) | < 0.0001 |
| GDP | -0.424 | (-0.76, -0.09) | 0.013 |
| Urbanization | -0.378 | (-1.05, 0.30) | 0.27 |
| Distance from source | 0.287 | (-0.03, 0.61) | 0.08 |
| Air transport | -0.058 | (-0.17, 0.05) | 0.30 |
| GHS detection score | -0.267 | (-0.66, 0.12) | 0.18 |

<sup>†</sup> The estimated coefficient of a variable in log form in the AFT model is the elasticity of survival time with respect to the variable.

Another AFT analysis with the HFI replaced by its constituents, personal and economic freedom was performed (Table 2). Personal freedom had the largest effect (coefficient = 1.8). Economic freedom had a lower effect (coefficient = -0.273) and was not significant (p = 0.71). A model including personal freedom instead of the HFI had the lowest AIC of all the models tested. Thus, personal freedom is the variable that has the most influence on survival time. Cox proportional hazards analysis was also performed. Variable coefficients had similar values. However, the proportional hazards assumption was not met.

**Table 2 -.** AFT model with Human Freedom Index decomposed.

| **Variables** | **Coefficient<sup>†</sup>** | **95 % C.I.** | **P-value** |
| --- | --- | --- | --- |
| Personal freedom | -1.80 | (-2.94, -0.65) | 0.002 |
| Economic freedom | -0.273 | (-1.74, 1.19) | 0.71 |
| Population | -0.347 | (-0.51, -0.19) | < 0.0001 |
| GDP | -0.436 | (-0.77, -0.11) | 0.01 |
| Urbanization | -0.378 | (-1.00, 0.34) | 0.33 |
| Distance from source | 0.330 | (0.007, 0.65) | 0.045 |
| Air transport | -0.071 | (-0.18, 0.04) | 0.22 |
| GHS detection score | -0.224 | (-0.62, 0.17) | 0.26 |

<sup>†</sup> The estimated coefficient of a variable in log form in the AFT model is the elasticity of survival time with respect to the variable.

## 4. Discussion

This study suggests that some of the factors associated with disease spread in the theoretical framework of the proposed gravity model are supported by empirical data from the current COVID-19 pandemic. Previous attempts to relate the gravity model to spread in the context of a pandemic used generalized linear models rather than survival analysis to model time to index case (Li et al., 2011). Past work on the topic also did not include potential confounders such as personal freedom, air travel volume and urbanization. Opportunities to conduct an ecological study of this type are as rare as major pandemics. Spread of the disease to the entire world allows for a survival analysis with more data points and no censoring, which leads to more precise estimates.

Selection bias is an important issue in ecological studies. Only countries that had experienced their index case by the 11^th^ of April were included. This represents the vast majority of countries in the world. However, generalizability of the study to the Pacific Island nations and other countries not included might be limited. The potential for information bias should also be brought up. Data upon which indices like the Global Health Security Index are constructed is self-reported by countries. Nonetheless, alternatives of comparable comprehensiveness are not available.

A key finding of the study is the fact that higher personal freedom is associated with less time until a pandemic reaches a country. Potential confounding could be caused by the tendency of countries with lower personal freedom to underreport and underdiagnose (Kavanagh, 2020). However, the Global Health Security Detection and Reporting score mitigates at least some of the confounding, as it accounts for the capacity and willingness of countries to report. This could have implications in the policy of the early stages of a pandemic. Most countries have opted for social distancing measures which reduce personal freedom, like limiting public gatherings and travel restrictions (Lewnard & Lo, 2020). Consensus on the effectiveness of these measures has not been reached. Countries like Sweden are trying to limit social and economic disruption, even though certain models predict high mortality associated with this strategy (Gardner et al., 2020).

Implementing restrictions as soon as a potentially pandemic virus starts spreading could delay its arrival to a country. If countries had more time to prepare their response, there would be a lower probability of straining health systems and thus fewer deaths. This type of analysis could be used to guide risk assessment and identify countries that are likely to be affected sooner in the course of a pandemic. These high-risk countries that have less time to spare would benefit from even more attention to pandemic preparedness. The results merit further investigation into the application of the model at the district and regional level, to assess whether it can be used at a smaller scale.

In conclusion, the gravity model-based survival analysis managed to measure the influence of important socio-economic variables on the time from the beginning of a pandemic to the first case in a country. Ecological survival analysis at the country level can aid in identifying patterns of spatial and temporal spread and potentially provide insight into the influence of social and economic factors on the global transmission of viral diseases.

## Data availability

The data that support the findings of this study are available on figshare at https://doi.org/10.6084/m9.figshare.12205265.v1. These data were derived from the public domain sources listed as references in the link and manuscript.

## References

Bozdogan, H. (1987). Model selection and Akaike’s Information Criterion (AIC): The general theory and its analytical extensions. Psychometrika, 52(3), 345–370. https://doi.org/10.1007/BF02294361

[dataset] Cato Institute. (2019). The Human Freedom Index 2019. https://www.cato.org/sites/cato.org/files/human-freedom-index-files/cato-human-freedom-index-update-3.pdf

[dataset] ECDC. (2020a, April 11). COVID-19 Coronavirus data. https://data.europa.eu/euodp/en/data/dataset/covid-19-coronavirus-data

[dataset] ECDC. (2020b, April 23). Situation update worldwide. https://www.ecdc.europa.eu/en/geographical-distribution-2019-ncov-cases

Eisenberg, J. N. S., Desai, M. A., Levy, K., Bates, S. J., Liang, S., Naumoff, K., & Scott, J. C. (2007). Environmental Determinants of Infectious Disease: A Framework for Tracking Causal Links and Guiding Public Health Research. Environmental Health Perspectives, 115(8), 1216–1223. https://doi.org/10.1289/ehp.9806

Gardner, J. M., Willem, L., van der Wijngaart, W., Kamerlin, S. C. L., Brusselaers, N., & Kasson, P. (2020). Intervention strategies against COVID-19 and their estimated impact on Swedish healthcare capacity [Preprint]. Epidemiology. https://doi.org/10.1101/2020.04.11.20062133

Grambsch, P. M., & Therneau, T. M. (1994). Proportional hazards tests and diagnostics based on weighted residuals. Biometrika, 81(3), 515–526. https://doi.org/10.1093/biomet/8L3.515

[dataset] Johns Hopkins Center for Health Security, N. T. I. (2019). Global Health Security Index. https://www.ghsindex.org/wp-content/uploads/2020/04/2019-Global-Health-Security-Index.pdf

Kavanagh, M. M. (2020). Authoritarianism, outbreaks, and information politics. The Lancet Public Health, 5(3), e135-e136. https://doi.org/10.1016/S2468-2667(20)30030-X

Kraemer, M. U. G., Golding, N., Bisanzio, D., Bhatt, S., Pigott, D. M., Ray, S. E., Brady, O. J., Brownstein, J. S., Faria, N. R., Cummings, D. A. T., Pybus, O. G., Smith, D. L., Tatem, A. J., Hay, S. I., & Reiner, R. C. (2019). Utilizing general human movement models to predict the spread of emerging infectious diseases in resource poor settings. Scientific Reports, 9(1), 5151. https://doi.org/10.1038/s41598-019-41192-3

Lewnard, J. A., & Lo, N. C. (2020). Scientific and ethical basis for social-distancing interventions against COVID-19. The Lancet Infectious Diseases, S1473309920301900. https://doi.org/10.1016/S1473-3099(20)30190-0

Li, X., Tian, H., Lai, D., & Zhang, Z. (2011). Validation of the Gravity Model in Predicting the Global Spread of Influenza. International Journal of Environmental Research and Public Health, 8(8), 3134–3143. https://doi.org/10.3390/ijerph8083134

Mangili, A., Vindenes, T., & Gendreau, M. (2015). Infectious Risks of Air Travel. Microbiology Spectrum, 3(5). https://doi.org/10.1128/microbiolspec.IOL5-0009-2015

Nay, O. (2020). Can a virus undermine human rights? The Lancet Public Health, S246826672030092X. https://doi.org/10.1016/S2468-2667(20)30092-X

Robert J. Hijmans. (2019). Package ‘geosphere’. https://cran.r-project.org/web/packages/geosphere/geosphere.pdf

Rothan, H. A., & Byrareddy, S. N. (2020). The epidemiology and pathogenesis of coronavirus disease (COVID-19) outbreak. Journal of Autoimmunity, 109, 102433. https://doi.org/10.10167j.jaut.2020.102433

Tatem, A. J., Rogers, D. J., & Hay, S. I. (2006). Global Transport Networks and Infectious Disease Spread. In Advances in Parasitology (Vol. 62, pp. 293–343). Elsevier. https://doi.org/10.1016/S0065-308X(05)62009-X

Viboud, C., Bjornstad, O. N., Smith, D. L., Simonsen, L., Miller, M. A., & Grenfell, B. T. (2006). Synchrony, Waves, and Spatial Hierarchies in the Spread of Influenza. Science, 312(5772), 447–451. https://doi.org/10.1126/science.1125237

Wei, L. J. (1992). The accelerated failure time model: A useful alternative to the cox regression model in survival analysis. Statistics in Medicine, 11(14-15), 1871–1879. https://doi.org/10.1002/sim.4780111409

[dataset] World Bank. (n.d.). World Bank Open Data. https://data.worldbank.org/

